# Exposure to Renin-Angiotensin System Inhibitors Is Associated with Reduced Mortality of Older Hypertensive Covid-19 Patients

**DOI:** 10.1101/2020.12.15.20247999

**Authors:** Mauro Gori, Carlo Berzuini, Emilia D’Elia, Arianna Ghirardi, Luisa Bernardinelli, Antonello Gavazzi, Giulio Balestrieri, Andrea Giammarresi, Roberto Trevisan, Fabiano Di Marco, Antonio Bellasi, Mariangela Amoroso, Federico Raimondi, Luca Novelli, Bianca Magro, Gianpaolo Mangia, Ferdinando L. Lorini, Giulio Guagliumi, Stefano Fagiuoli, Gianfranco Parati, Michele Senni

**Author notes:** Corresponding author:* Gianfranco Parati, MD, FESC, Dept. Cardiovascular, Neural and Metabolic Sciences, San Luca Hospital, IRCCS, Istituto Auxologico Italiano & University of Milano-Bicocca; P.za Brescia, 20; 20149 Milan, Italy. First coauthors equally contributing to this work (Mauro Gori, Carlo Berzuini). Last coauthors equally contributing to this work (Gianfranco Parati, Michele Senni).

## Abstract

From a cohort of 1352 consecutive patients admitted with coronavirus disease (Covid-19) to Papa Giovanni XXIII Hospital in Bergamo, Italy, between February and April 2020, we selected and studied 688 patients with arterial hypertension (254 deaths) to assess whether use of renin-angiotensin system inhibitors (RASIs) prior to hospital admission affects mortality from Covid-19. Prior use of RASIs was associated with a lower mortality in the over-68 group of patients, whereas no evidence of a similar effect (whether protective or adverse) was found in the younger group. There was positive relative excess due to a statistically significant (*p* =0.001) interaction between prior RASI exposure and an age greater than 68 years, corresponding to a positive relative excess risk. Next we used the subgroup of 411 hypertensive patients older than 68 yrs to separately assess the effects prior use of two RASI drug subclasses, angiotensin-converting enzyme inhibitors (ACEIs) and angiogiotensin receptor blockers (ARBs), by comparing these two exposures with no exposure to RASIs. We found both prior use of ACEIs and prior use of ARBs to be associated with a lower Covid-19 mortality, after adjusting for 32 medical history variables via propensity score matching. (*OR*_*ACEI*_ = 0.57, 95%CI 0.36 to 0.91, *p* =0.018) (*OR*_*ARB*_ = 0.49, 95%CI 0.29 to 0.82, *p* =0.006).

## Introduction

Of interest here is the impact of renin-angiotensin system (RAS) inhibitor drugs on the consequences of coronavirus disease (Covid-19). RAS inhibitor drugs (RASIs) represent a mainstay treatment for hypertension certain cardiological conditions involved in a severe course of Covid-19.

A possible adverse effect of RASIs in Covid-19 patients is suggested by the following biological argument. A negative regulator of RAS, called the Angiotensin I Converting Enzyme 2 (ACE2), has been found to act as a receptor for the Covid-responsible virus (SARS-Cov-2) to enter the infected’s cells and replicate^1^. Evidence from animal studies suggests that certain RASIs might upregulate ACE2^2^ and, as a consequence, help SARS-Cov-2 invade human cells. Most popular RASIs are angiotensin-converting enzyme inhibitors (ACEIs) and angiotensin receptor blockers (ARBs), both of which are widely used in the most Covid-19-vulnerable part of the population: the elderly. The prospect of a Covid-19-vulnerable population making widespread use of drugs suspected to worsen that disease cannot but raise deep concern.

These fears have been tempered by recent epidemiological studies showing no evidence of whatsoever effect of RASIs on Covid-19 outcomes^3^.

Our study changes this picture by opening a new perspective on the role of RASIs in Covid-19. The study is based on 1352 consecutive Covid-19 patients admitted to Papa Giovanni XXIII Hospital in Bergamo between February 23rd and April 7th, 2020 (median follow-up 34 days), changes this picture.

We found that use of ARBs or ACEIs for a certain period of time up to hospital admission is associated with a lower mortality among elderly hypertensive Covid-19 patients, after adjusting for medical history, whereas no evidence of a RASIs effect was found in the younger Covid-affected hypertensive populatio n. Effects of ACEIs and ARBS were assessed separately.

This scientific advance has been possible thanks to a correct use of data analysis methods. In contrast with most previous studies in this area, we have recognized/assessed the striking difference between the effect of RASI exposure in the young vs the old Covid-19 populations, and allowed our effect estimates to vary between age strata, for a reliable estimate of the effect of RASIs in the older, and most vulnerable, stratum of the Covid-19 population, with implications for public health intervention and design of future studies in this area.

The above argument is splendidly illustrated by a study by Fosboel and colleagues.^4^ In common with our study, these authors investigate the effect of prior use of RASIs in hypertensive Covid-19 patients. They obtain an adjusted hazard of death ratio estimate of 0.83 with a 95% confidence interval (0.67-1.03) that barely covers the null and a *p*-value of .09. Their conclusion that “prior use of ACEI/ARBs was not significantly associated with COVID-19 diagnosis or mortality among patients with hypertension” is rash. Had they taken age-related effect modification into account, thereby avoiding effect dilution due to inclusion of young patients, they would have probably seen evidence of a statistically significant Covid-19-beneficial effect of ACEI/ARBs in the older age.

## Methods

### Ethics

Necessary approval was obtained from the Bergamo Ethics Committee (n. 37/2020) with operating center at the Papa Giovanni XXIII Hospital of Bergamo, which has capacity to give its opinion on studies conducted in a number of health care structures in the Bergamo area of the north-italian region Lom bardia, including the mentioned hospital, ten “Aziende Socio Sanitarie Territoriali” and fourteen “Agenzie di Tutela della Salute”. In conformity with local protocol, consent was obtained from the patient.

### Data

We included in our study all patients older than 18 years with positive rhino-pharyngeal swab for SARS-CoV-2 infection, hospitalized for Covid-19 at Papa Giovanni XXIII Hospital (a tertiary hospital of 1080 beds located in Bergamo, the initial epicenter of Italian Covid-19 storm), between February 23rd and April 7th, 2020. Patient follow-up ended on May 5th, 2020. Follow-up time had a median of 34 days and an Inter Quartile Range (IQR) of 19 to 41.

Our initial sample included 1352 consecutive Covid-19 patients, diagnosed on the basis of the updated WHO interim guidance^5^. Information about the history and physical examination of patients hospitalized with Covid-19 was derived via chart review by medical officers. Variables collected through standardized recording forms included age, sex, comorbidities, dates of symptoms’ onset and hospital admission. Hypertension was defined as having a diastolic blood pressure equal or greater than 90 mm Hg and/or a systolic blood pressure equal to or greater than 140 mmHg and/or a history of antihypertensive medication use. Laboratory confirmation of SARS-CoV-2 infection via SARS-CoV-2 genome detection from nasal swab and respiratory samples was obtained through two different molecular methods (GeneFinder COVID-19-Elitech Group, Allplex™2019-nCoV Assay-Seegene Inc) following instructions. After purification of viral RNA from clinical samples, presence of RdRp, E and N viral genes was detected by using real time Polymerase Chain Reaction (RT-PCR) according to WHO protocol.

Data were subjected to quality checks, validated for internal consistency and then anonymized prior to transfer.

### Outcome

Primary endpoint was mortality from all-causes, either occurring in-hospital or shortly after discharge.

### Statistical analysis

No sample-size calculations were performed. Age was dichotomized via median split as *≤* 68 or *>*68yrs, with no attempt to optimize the divide^6^. We shall hereafter use the symbol 68+ as a shorthand for *>*68yrs. Analysis in this paper focuses on hypertensive patients. Our exposure groups were: prior RASIs-users, no-RASIs-use, prior ACEIs-users and prior ARBs-users. Pairwise comparisons between these groups^7^ in terms of mortality from Covid-19 were performed by using propensity score matching methods^8^ to adjust for potential pre-hospitalisation confounders and logistic regression of the binary survival outcome on the exposure variable.^9,10^

We acknowledged apossible modification of RASIs effect due to age by allowing our model of survival outcome to include an additional term for a possible interaction between the effect of RASIs and that of age. A significantly different value of this parameter from zero would represent evidence of the two effects interacting on a multiplicative scale. Because interaction is more relevant to public health if expressed on an additive scale^11^, this paper presents evidence of `RASI` *×* `age` interaction also in a relative excess risk (RER) form^12^ after appropriate dichotomisation^13^ of the continuous age variable (*≤*68 vs 68+). A positive RER is obtained where there is a “target” age stratum where a real-world intervention in favour of RASIs is likely to have greater impact than in remaining population.

Analyses were conducted with `R` software by exploiting the `R Studio` user interface^14^.

## Results

### Patients

Our initial sample included 1352 patients. There were 353 (26.1%) deaths. Table 1 summarizes demographic, home therapy and comorbidity data of our patients.

**Table 1.**
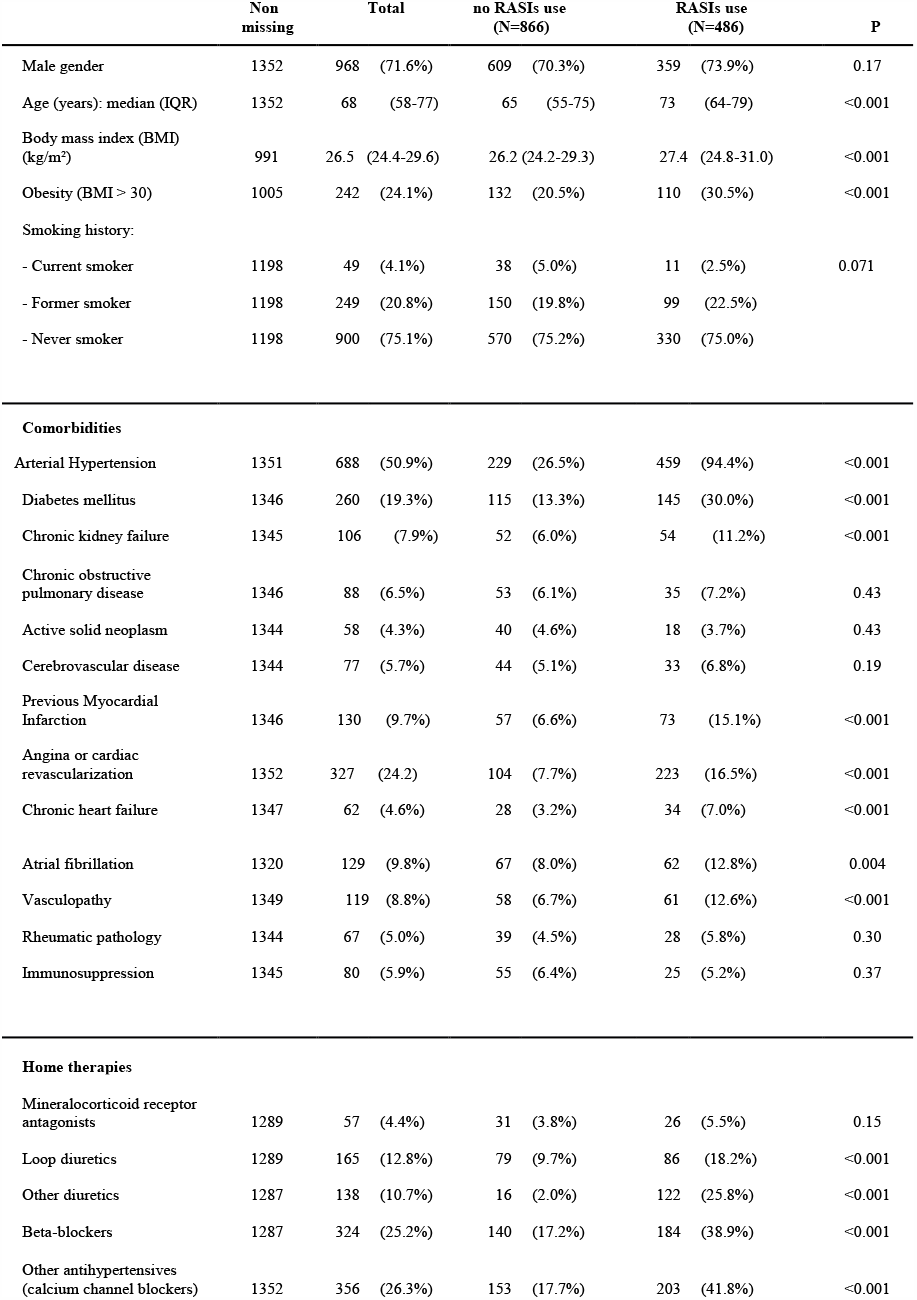
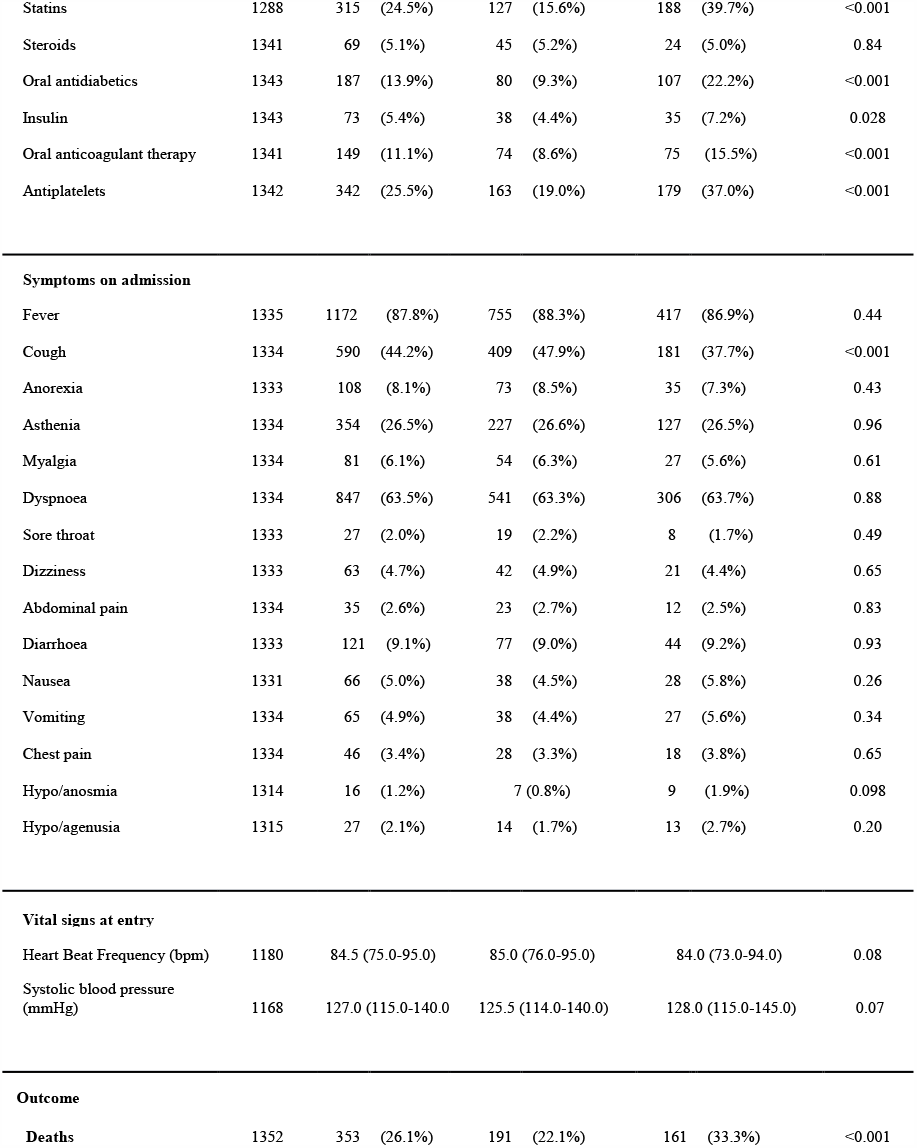
Characteristics of the global sample of patients are here reported by stratifying by RASIs-use vs no-RASIs-use. Symbol N stands for group numerosity. Symbol P stands for p-value for the difference between RASIs-use and no-RASIs-use populations with respect to a specific characteristic. For each yes-no characteristic (eg., male gender) the table reports number and percentage of “yes” patients within a particular stratum.

Missing values were imputed via `R` package `MICE`. No signs of systematic missingness were detected. `Obesity` and `Smoking` were excluded from analysis due to a percentage of missing values in excess of 5%. Results from a subsidiary analysis restricted to the set of patients with complete information about these two variables, and performed by including these in the models, did not yield appreciably different results from the main analysis.

Table 1 compares the 486 patients on RASIs at admission with the remaining 866. A total of 968 patients (71.6%) were men; median age was 68 years (IQR: 58 to 77). RASI users tended to be older (median age: 73 years vs 65 years) and had an overall higher prevalence of comorbidities, eg. hypertension (94.4% vs 26.5%), diabetes (30.0% vs 13.3%), chronic kidney disease (11.2% vs 6.0%), previous myocardial infarction (15.1% vs 6.6%), chronic heart failure (7.0% vs 3.2%), angina or previous cardiac revascularization (16.5% vs 7.7%), atrial fibrillation (12.8% vs 8.0%) and vasculopathy (12.6% vs 6.7%). They were also more frequently treated with diuretics, beta-blockers, calcium channel blockers, statins, anti-diabetics and antiplatelet drugs.

Table 2 illuminates a main point. As our attention shifts from the whole population to the smaller stratum of hypertensives and then, from within this stratum, to that of *older hypertensives*, a protective effect of RASIs progressively emerges from a simple scrutiny of raw counts. (Whole population: RASI 32% vs no-RASI 24%; hypertensives of all ages: RASI 32% vs no-RASI 40%; 68+ hypertensives: RASI 40% vs no-RASI 58%).

**Table 2.** Size and mortality are reported for each stratum by age, hypertension indicator, survival status and RASI-exposure indicator.

|  | whole population |  | hypertensives all ages |  | hypertensives 68+ |  |
| --- | --- | --- | --- | --- | --- | --- |
|  |  | decedents |  | decedents |  | decedents |
| Exposure | surv | (mortality) | surv | (mortality) | surv | (mortality) |
| no RASI | 690 | 221 (24%) | 154 | 106 (40%) | 63 | 89 (58%) |
| RASI | 324 | 155 (32%) | 304 | 148 (32%) | 153 | 106 (40%) |

### MAIN RESULT 1: Age modifies RASI effect on Covid-19 outcome for hypertensive patients

The variables in Table 1 (except for the survival outcome) were considered for construction of a propensity score model for chronic use of RASIs. The model parameter estimates were used to calculate RASI-propensity score for each sample patient. Two subgroups of patients, RASI-exposed vs RASI-free, were created by matching them with respect to the score. Based on the union of these two groups, we performed a logistic regression of the binary survival outcome on the RASI-exposure indicator, also allowing this indicator and the binary (*<*68 vs 68+) age indicator to interact in their effects on the outcome. Effect of continuous age was modelled non-parametrically via splines. There was significant (*p*=0.001) evidence of interaction, corresponding to a RER of 0.19, signifying that older Covid-19 patients with hypertension gain more from prior exposure to RASIs (OR=0.5, *p*=0.07) than their younger “colleagues”.

### MAIN RESULT 2: In an analysis of 68+ hypertensive Covid-19 patients, both pre-hospital exposure to ARBs and pre-hospital exposure to ACEIs were associated with a lower mortality, when compared with no exposure to RASIs. Effect was statistically more significant in the ARB group. 8 patients with prior exposure to both ARBs and ACEIs were excluded

The above conclusion sare supported by the crude statistics of Table 3 and by the analysis described in the following, where adjustment for confounders is made.

**Table 3.** In this table, 68+ hypertensive Covid-19 patients are cross-stratified by outcome status (rows) and according as they do not use RASIs (column 2), or use ACEIs but not ARBs (column 3), or use ACEIs but not ARBs (column 4). Reported in brackets are stratum-specific crude mortality estimates. As we move from column 2 to 4, we encounter a progressively lower crude estimate of mortality.

| OUTCOME | RASI non-users (mortality) | prior use of ACEIs (mortality) | prior use of ARBs (mortality) |
| --- | --- | --- | --- |
| survived | 69 | 83 | 74 |
| decedents | 90 (56%) | 61 (42%) | 46 (38%) |

First we contrasted “prior ACEI users” to “no-RASI-users”, and then “prior ARB users” to “no-RASI-users”, each comparison involving a separate calculation of the appropriate propensity score (see Table 4 for the propensity score used in the ARBs vs no-RASIs comparison).

**Table 4:**
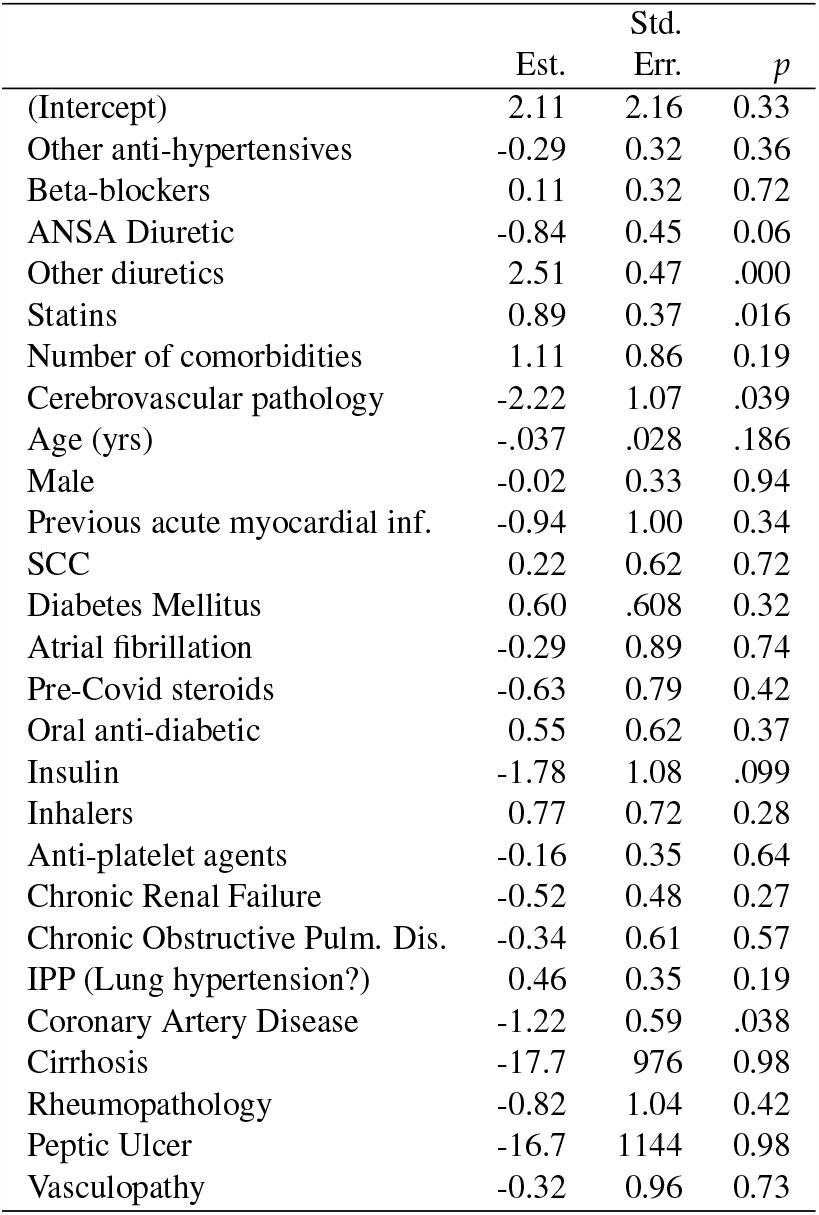
estimated propensity score for chronic use of ARB in population of 68+ Covid-19 patients with hypertension (ACEI users excluded). The score provides an empirical estimate of the probability of the generic 68+ hypertensive patient having been exposed to ARBs prior to hospitalization, conditional on their medical history and on not having used ACEIs. Std.

| | Est. | Std. Err. | $p$ |
| --- | --- | --- | --- |
| (Intercept) | 2.11 | 2.16 | 0.33 |
| Other anti-hypertensives | -0.29 | 0.32 | 0.36 |
| Beta-blockers | 0.11 | 0.32 | 0.72 |
| ANSA Diuretic | -0.84 | 0.45 | 0.06 |
| Other diuretics | 2.51 | 0.47 | .000 |
| Statins | 0.89 | 0.37 | .016 |
| Number of comorbidities | 1.11 | 0.86 | 0.19 |
| Cerebrovascular pathology | -2.22 | 1.07 | .039 |
| Age (yrs) | -.037 | .028 | .186 |
| Male | -0.02 | 0.33 | 0.94 |
| Previous acute myocardial inf. | -0.94 | 1.00 | 0.34 |
| SCC | 0.22 | 0.62 | 0.72 |
| Diabetes Mellitus | 0.60 | .608 | 0.32 |
| Atrial fibrillation | -0.29 | 0.89 | 0.74 |
| Pre-Covid steroids | -0.63 | 0.79 | 0.42 |
| Oral anti-diabetic | 0.55 | 0.62 | 0.37 |
| Insulin | -1.78 | 1.08 | .099 |
| Inhalers | 0.77 | 0.72 | 0.28 |
| Anti-platelet agents | -0.16 | 0.35 | 0.64 |
| Chronic Renal Failure | -0.52 | 0.48 | 0.27 |
| Chronic Obstructive Pulm. Dis. | -0.34 | 0.61 | 0.57 |
| IPP (Lung hypertension?) | 0.46 | 0.35 | 0.19 |
| Coronary Artery Disease | -1.22 | 0.59 | .038 |
| Cirrhosis | -17.7 | 976 | 0.98 |
| Rheumopathology | -0.82 | 1.04 | 0.42 |
| Peptic Ulcer | -16.7 | 1144 | 0.98 |
| Vasculopathy | -0.32 | 0.96 | 0.73 |

Results in Table 5 can be summarised as follows:

**Table 5.**
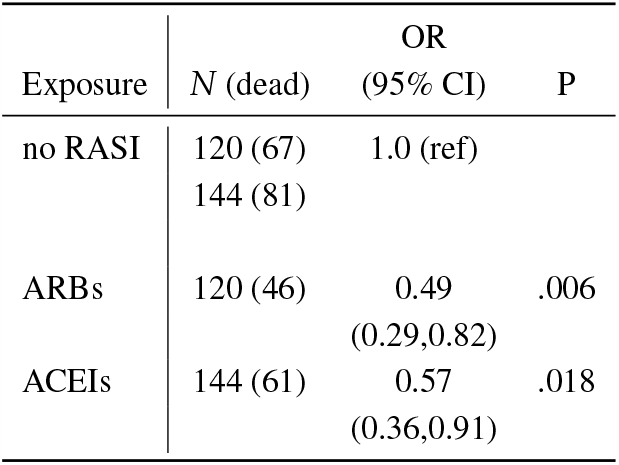
This table presents ORs for the ARB/no-ACEI and the ARB/no-ACEI exposure groups (within the population of 68+ hypertensives), expressed relative to the RASI-free group (which has been chosen as reference). All the ORs are adjusted for pre-hospitalization variables via propensity-score matching, as described in the main text of this paper. Point estimate for the ORs are accompanied by their corresponding 95% confidence intervals, in brackets. These estimates are based on the reduced samples sizes produced by the matching (120 per exposure group in the assessment of ARB vs RASI-free effect; 144 per exposure group in the assessment of ARB vs RASI-free effect. Both effects appear to be significantly different from zero. ACEI effect appears lower in magnitude (higher OR) than that of ARBs, but not significantly so.

Chronic ACEIs use, when compared with no RASI-use within the stratum of 68+ hypertensives, was found to be significantly associated with a lower mortality (P = 0.018, OR= 0.57, 95%CI 0.36 to 0.91), after adjusting for medical history.

Chronic ARBs use, when compared with no RASI-use within the stratum of 68+ hypertensives, was found to be significantly associated with an even lower mortality (P = 0.006, OR 0.49, 95%CI 0.29 to 0.82), after adjusting for variability induced by use of concomitant drugs and by pre-existing morbidities.

### Diagnostic Analyses

This section provides the Reader, with more insight into the part of our analysis that concerns the comparison between prior ARB-users and no-RASI-users. It looks into possible differences between these two exposure groups in terms of post-hospitalization variables.

Table 6 does not reveal marked differences between two matched ARB and no-RASI groups (120 patients each) in terms of clinical observations on hospital admission. These are the two groups used to assess the effect of ARBs in the population of 68+ hypertensives (the omitted variables showed less important discrepancies).

**Table 6.** This table compares the ARB/noACEI-exposed group of 120 matched patients with the corresponding group of 120 matched RASI-free patients (68+ hypertensives) in terms of some clinical variables measured upon hospital admission (those variables that revealed greater standardized discrepancies were chosen). There are no marked differences.

| Exposure | n | Fever | Cough | Dispnea | Respiratory Insufficiency |
| --- | --- | --- | --- | --- | --- |
| RASI free | 136 | 105 | 41 | 78 | 104 |
| ARBs | 136 | 120 | 40 | 84 | 106 |

|  | n | RX anomalies | pO<sub>2</sub> | pCO<sub>2</sub> | Respiratory FiO<sub>2</sub>max100 | Bilateral P/F |
| --- | --- | --- | --- | --- | --- | --- |
| RASI free | 136 | 107 | 71.7 | 31.9 | 0.46 | 192 |
| ARBs | 136 | 95 | 76.4 | 32.3 | 0.44 | 206 |

Nor do the sam e two groups markedly differ in terms of biochemical parameters measured upon hospital admission, according to Table 7. The omitted parameters showed even smaller standardized discrepancies).

**Table 7.** This table compares the same groups in the previous table in terms of some biochemical parameters measured upon hospital admission (those parameters that revealed greater standardized discrepancies were chosen). There were no marked differences.

| Exposure | n | LDH | AST | ALT | BILI | CREA | WBC | LYMPHco |
| --- | --- | --- | --- | --- | --- | --- | --- | --- |
| RASI-free | 136 | 467.79 | 64.41 | 45.66 | 0.73 | 1.16 | 8627 | 828 |
| ARBs | 136 | 520.57 | 93.97 | 72.84 | 0.72 | 1.62 | 8476 | 883 |

|  | n | Lymph% | NEUTco | NEUT% | PLT | PCR | UREA | HB |
| --- | --- | --- | --- | --- | --- | --- | --- | --- |
| RASI-free | 136 | 12.19 | 7121 | 82.1 | 216647 | 13.99 | 49.92 | 12.72 |
| ARBs | 136 | 11.89 | 7411 | 81.4 | 235448 | 12.72 | 58.22 | 12.54 |

Nor do the same two groups markedly differ in terms of age, comorbidities and chronic therapies, according to Table 8.

**Table 8:** this table compares propensity-matched exposure groups (ARB/no-RASI vs RASI-free) in terms of comorbidity frequencies, frequencies of home therapies and average age.

| Exposure | n | Mean num of comorbid. | ANSA diuretics | Other diuretics | Cereb. vasc. path. |
| --- | --- | --- | --- | --- | --- |
| RASI-free | 136 | 0.86 | 31 | 37 | 7 |
| ARBs/no ACEIs | 136 | 0.83 | 27 | 47 | 9 |

| ARBs? | Statins | Insulin | Inhalers | Chronic Obstr. Pulm. Disease | Age |
| --- | --- | --- | --- | --- | --- |
| no | 63 | 10 | 9 | 24 | 78.0 |
| yes | 65 | 13 | 9 | 22 | 78.0 |

Prior ARBs users and no-RASI users had no significantly different chance of Intensive Care Unit (ICU) assignment, or of assignment to specific in-hospital treatments, after adjusting for medical history (ICU: OR= 1.022, *p* = 0.45; Tocilizumab: OR = 0.99, *p* = 0.32; Antibiotics: OR = 0.97, *p* = 0.68; Steroids: OR = 1.00, *p* = 0.73; Hydroxychloroquine: OR 0.963, *p* = 0.47; Kaletra: OR = 0.97, *p* = 0.56; Oseltamivir: OR = 0.985, *p* = 0.15). Nor were these two groups significantly different in terms of clinical picture.

Effectiveness of the matching in our analysis of ARBs effect can be visually appreciated in Figure 1. For an explanation of the figure we refer the Reader to its legend.

**Figure 1.**
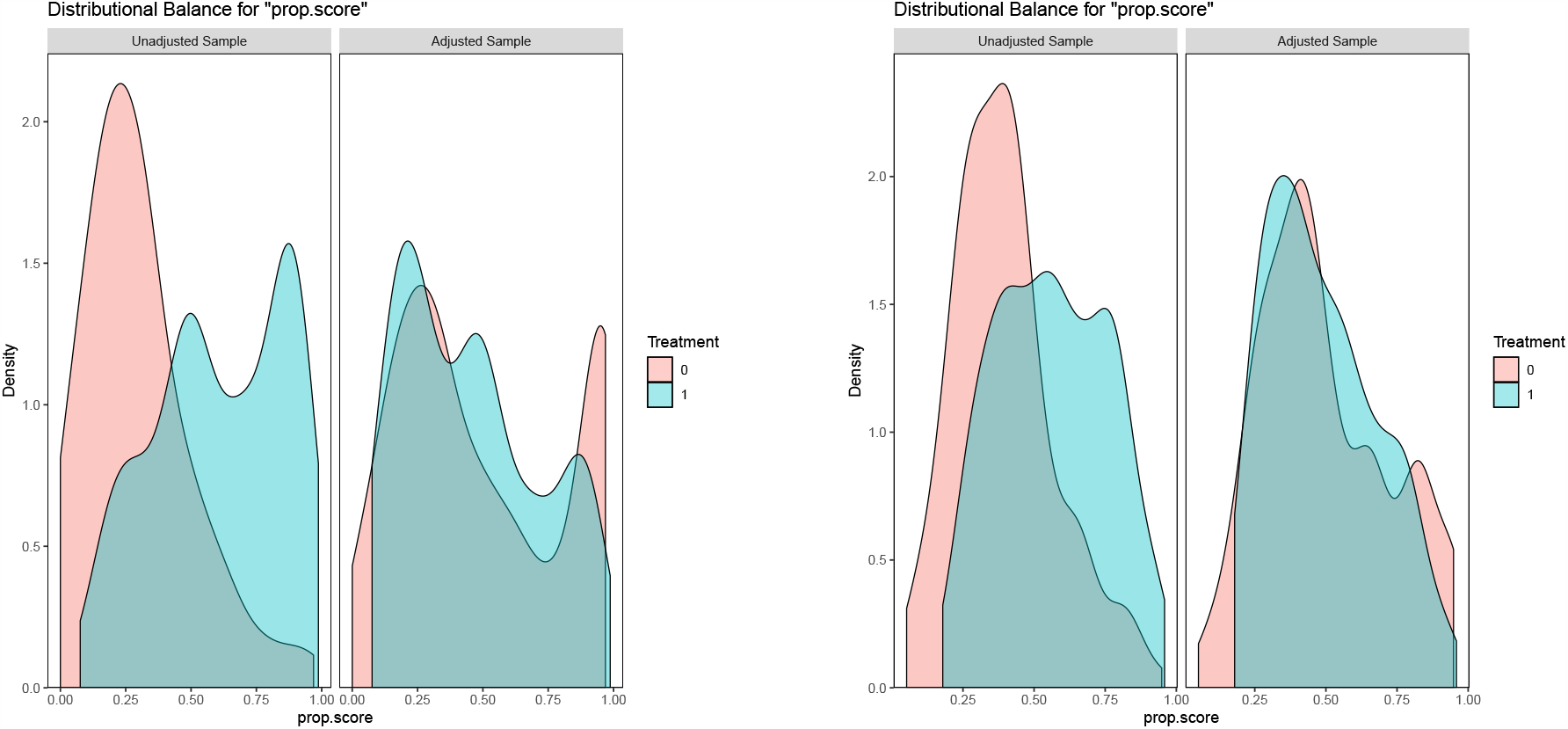
This figure allows a visual assessment of the degree of balancing achieved by the matching in our analyses of the effects of ARBs (two plots in the left half of the figure) and of ACEIs (two plots in the right half) on mortality in the population of 68+ Covid-19 patients with arterial hypertension. The figure contains four plots. Moving from left to right, plots 1 and 2 shows the estimated densities of the propensity score for ARB use within the groups of ARB users (blue) and non-RASI users (pink), before (plot 1) and after (plot 2) the matching. The degree to which the densities overlap is a good measure of group comparability. Hence the good overlap of our matched “ARB” and “no-RASI” groups in plot 2 reassures us that a statistically significant difference in mortality between our matched “ARB” and “no-RASI” will represent unbiased evidence of an effect of chronic use of RASIs on mortality in the studied population. Note from Plot 1 the unmatched exposure groups were *not* comparable. We conclude that the matching has played a crucial role in creating conditions for a credible estimate of ARBs effect. Completely analogous remarks can be made on the basis of plots 3 and 4 in relation with our assessment of the effect of pre-hospitalization exposure to ACEI on mortality in the population of 68+ Covid-19 patients with arterial hypertension.

### Effect of post-admission RASIs discontinuation

As show n in Tab le 9, we had 484 chronic 68+ RASI users with with hypertension. Of these, 138 remained on RASIs after hospitalization. ACEI users were slightly more numerous than ARB users, and they exhibited a higher rate of post-hospitalization drug continuation. According to Table 10, crude mortality was higher in patients who had RASIs discontinued, when compared to those who remained on RASIs.

**Table 9:** Our sample of chronic RASIs users stratified by drug class and post-admission continuation vs discontinuation of the drug.

| Group | discontinued | continued | continuation rate |
| --- | --- | --- | --- |
| ARBs users | 173 | 59 | 25.4% |
| ACEi users | 182 | 79 | 30% |

**Table 10:** statistics within our sample of 68+ hypertensive patients. According to results in this table, mortality from Covid-19 is higher in the group of sample patients who discontinued use of RASI at hospital admission, as compared to patients who continued it, and in both these groups mortality is lower than in no-RASI users.

| Outcome | no RASIs users | RASIs discontinued | RASIs continued |
| --- | --- | --- | --- |
| survived | 63 | 105 | 57 |
| decedents | 89 | 86 | 25 |

We did not further examine effect of continuation as we felt we had no complete record of all the information that may have influenced the hospital doctor’s decision to continue/discontinue RASIs (see Discussion).

## Discussion

Our study finds that elderly hypertensive Covid-19 patients using ARBs or ACEIs before hospital admission have a significantly lower mortality than sim ilar patients who did not use RASIs, after adjusting for the patien t’ s medical history. It finds no evidence of a RASIs effect in similar, but younger, patients.

The protective effect found in the elderly can be explained by the ability of RASIs to avert Covid-induced cardiovascular complications^15^.

Our study shows that Covid-19 lethality increases with age and in the presence of comorbidities (eg. hypertension) associated with higher vulnerability to the cardiorespiratory complications of Covid-19^16^, and that RASIs may lower mortality of Covid-19 patients with hypertension or other cardiovascular comorbidities. Protective effect of RASIs may take place through antagonism of the deleterious effects of Ang II. Liu and colleagues^17^ report serum Ang II plasma levels in a sample of twelve Covid-19 infected patients as being markedly elevated and linearly associated with viral load and lung injury. These findings support the hypothesis that elevated levels of Ang II may foster acute respiratory distress syndrome (ARDS) in Covid-19 patients, which would explain the protective role of RASIs found in older Covid-19 patients. Additionally, in vitro cells treatment with Ang II was found to enhance ACE2 ubiquitination also mediated by AT1R, ultimately stimulating ACE2 lysosomal degradation^18^. This might prevent interaction of SARS-CoV-2 with ACE2 catalytic site. Noteworthily ARBs, through AT1R antagonism, have been suggested as drugs able to prevent virus/ACE2 interaction, such pathway representing a putative mechanism by which ARBs, more than ACEIs, might prevent SARS-CoV-2 cells entry^19^. Indeed, our results point in this direction.

It could be argued that beneficial effect of previous RASIs use, especially of ARBs, is related to higher ACE2 expression with aging^20^. Thus, the older the patient the higher might ACE2 expression be and, concordantly, the greater might the RASIs beneficial effect in Covid-19 be. Finally, RASIs have antithrombotic properties that could further ameliorate the clinical course of Covid-19, possibly by reducing the thromboembolic complications associated with this disease^21^.

Unlike previous works, the present study avoids a serious methodological missteps by taking a correct approach to effect modification due to age, in such a way to avoid effect estimate dilution. Such a misstep might be a reason why results from a number of previous studies point in the same direction as ours without achieving nominal statistical significance, a notable example being the study by Fosboel and colleagues^22^.

The large, population-based study by Mancia and colleagues^23^ finds no evidence that ACEIs or ARBs affect risk of Covid-19. This result does not exclude ours, and for more than one reason. First, little detail is given in that paper about the “multivariable adjustment” used to calculate the effects of interest. The study relies on administrative data from regional databases with possibly incomplete information on comorbidities and drug use. More importantly, the outcome in Mancia’s study is *diagnosis* of severe Covid-19, rather than mortality (see Table 4 of the cited paper). In fact, of the four possible “coexisting conditions” that characterize individuals in their analysis (“respiratory disease”, “cardiovascular disease”, “kidney disease” and “cancer”), only the first turns out to to be characterized by a significant effect according to that study.

Our study was possible thanks to the fact that RASI drug allocation in the general population did not follow a fixed and uniform protocol based on the individual’s characteristics. In other words, primary care RASI prescription was relatively liberal. This is reflected by Figure 1, which shows that for each ARB-exposed patient we could find a patient with similar propensity score who had not used ARBs, for balanced comparison of the two treatment groups.

An open questionis whether the protective effects of RASIs are to be ascribed to their use *before* or to their use *after* the individual becomes infected. Or perhaps to both timings. Resolving this argument is beyond the scope of the present work. Under an observational regime, post-hospitalization administration of RASIs will be associated to prior exposure to the same drug and it will depend on decisions involving unrecorded information. Some authors concentrate on the effect of prior use of RASIs on patient admission parameters that appear to predict a severe outcome, under the (hard to test) assumption that those are *causal* parameters. Inference about the effect of post-admission therapeutic decisions should, ideally, be made via randomized studies, although the following example illustrates difficulties encountered by this approach. Just one problem being an ethical objection to randomizing assignment of a drug when an observational study has shown that the drug is likely to be beneficial.

Nevertheless, the above considerations represent a strong motivation to conduct a randomized clinical trial (RCT) to assess the effect of continuing/discontinuing RASIs in a patient hospitalized for Covid-19. One such trial has been performed: the BRACE CORONA trial^24^ assesses the effect of *discontinuing RASIs* on Covid-19 outcome. One problem with RCTs in a climate of health care urgency is that they require time. In order to circumvent this problem some studies fix a short follow-up horizon: 30 days in BRACE CORONA. Such a short time span may work only with a cohort of individuals at very high risk of severe outcome, which was not the case in BRACE CORONA. In fact BRACE CORONA, with a mortality rate of only 2.7%, records only 9 deaths per study arm. In spite of the low number of events, results from BRACE CORONA show a tendency towards a survival advantage of ACEIs/ARBs use. In the light of results from our study, it is not unreasonable to conjecture that had BRACE CORONA restricted admission to an old age stratum, or more in general to high risk patients, their results would have been in accord with ours. Other consideration: it is difficult to imagine an RCT where the randomized exposure is *prior* or *chronic* use of a drug. In fact, BRACE CORONA conditions on the patients having made chronic use of RASIs, and randomizes them over temporary suspension of the drug. Clearly the question addressed by BRACE CORONA is *not* equivalent to asking about the effect of chronic exposure, which is an interesting question in consideration of the possibility of these drugs acting via gradual and persisting structural changes.

By providing evidence of a beneficial effect of both ARBs and ACEIs, our study may be taken as suggesting that future RCTs should shine a light on both these drug classes.

### Strength and Limitations

Our study was based on a single big hospital. Rather than a limitation, this may be an element of strength of the study, insofar as homogeneity of target population reduces potential biases.

Two characteristics of our cohort, high percentage of elderly hypertensives and peak Covid-19 lethality, enhanced our power to detect the effects of interest.

We have used propensity-score matching methods to create exposure comparison groups that are comparable with respect to *observed* potential confounders. Despite the rather large number of medical history variables involved in the construction of our propensity scores, there may be additional unmeasured confounders that have not been taken into account, and consequently affect our results.

## Data Availability

Data may be requested to the Research Foundation of Bergamo Hospital, Bergamo, Italy (CovidLab)

## Funding

none.

### Author disclosures

*MG:* consulting fees from Novartis, Boehringer, and Menarini

*RT:* received lecture fees from AstraZeneca, Boehringer Ingelheim, Eli Lilly, Novo Nordisk, and Sanofi, and has participated in advisory panels for AstraZeneca, Boehringer Ingelheim, Eli Lilly, Novo Nordisk, and Sanofi.

*SF:* : speaker’s bureau and advisory board for Gilead, Abbvie, MSD, Novartis, Astellas, Intercept, Kedrion, Bayer.

*MS:* Bayer, Novartis, Boehringer, Astrazeneca.

*REMAINING AUTHORS:* none

## Author contributions

*MG, ED:* literature search, study design, data collection, writing.

*CB, LB:* statistical design of study, data analysis, writing and design of present document, literature search

*AG:* data analysis, supervision, writing, literature search, study design

*AG, GP, MS, SF, FLL, GG, FDM, RT:* supervision, writing, literature search, study design

*GB, AG, AB, MA, FR, LN, BM, GM:* data collection

## Footnotes

1 Markus Hoffmann et al. SARS-CoV-2 cell entry depends on ACE2 and TMPRSS2 and is blocked by a clinically proven protease inhibitor. *Cell*, 181(2):271–280.e8, 2020

2 G. O’Mara. Could ACE inhibitors and particularly ARBs increase susceptibility to COVID-19 infection? *British Medical Journal*, 2020

3 R Fosboel et al. Association of angiotensin-converting enzyme inhibitor or angiotensin receptor blocker use with COVID-19 diagnosis and mortality. *JAMA*, 2020; G Mancia, F Rea, M Ludergnani, G Apolone, and G. Corrao. Renin-Angiotensin-Aldosterone System Blockers and the Risk of Covid-19. *N Engl J Med*., 2020; J Li, X Wang, J Chen, H Zhang, and A Deng. Association of Renin-Angiotensin System Inhibitors With Severity or Risk of Death in Patients With Hypertension Hospitalized for Coronavirus Disease 2019 (COVID-19) Infection in Wuhan, China. *JAMA Cardiol*., 2020; C Gao, Y Cai, K Zhang, et al. Association of hypertension and antihypertensive treatment with COVID-19 mortality: a retrospective observational study. *Eur Heart J*.; H.R. Reynolds et al. Renin–angiotensin–aldosterone system inhibitors and risk of covid-19; and R Khera et al. Association of Angiotensin-Converting Enzyme Inhibitors and Angiotensin Receptor Blockers with the Risk of Hospitalization and Death in Hypertensive Patients with Coronavirus Disease-19. *medRxiv* : *the preprint server for health sciences*, 2020

4 R Fosboel et al. Association of angiotensin-converting enzyme inhibitor or angiotensin receptor blocker use with COVID-19 diagnosis and mortality. *JAMA*, 2020

5 https://www.who.int/publications-detail/laboratory-testing-for-2019-novel-coronavirus-in-suspectedhuman-cases-20200117

6 In order to avoid Type I error inflation, we used median age as the threshold without attempting to optimise it.

7 In order to avoid excessive test multiplicity, and consequent loss of power, not all the possible exposure comparisons were taken into consideration.

8 Paul R. Rosenbaum and Donald B. Rubin. The central role of the propensity score in observational studies for causal effects. *Biometrika*, 70(1):41–55, 04 1983

9 In accord with principles of causal inference there was no adjustment for post-hospitalization variables.

10 Carlo Berzuini, Alexander Philip Dawid, and Luisa Bernardinelli. *Causality: Statistical Perspectives and Applications*, volume 1 of *Wiley Series in Probability and Statistics*. John Wiley and Sons Ltd, Chichester, United Kingdom, July 2012

11 W. J. Blot and N. E. Day. Synergism and interaction: are they equivalent? *American Journal of Epidemiology*, 110:99–100, 1979; Rodolfo Saracci. Interaction and Synergism. *American Journal of Epidemiology*, 112(4):465–466, 10 1980; and Walker AM. Rothman KJ, Greenland S. Concepts of interaction. *Am J Epidemiol*., 112:467, 1980

12 Kenneth J. Rothman. CAUSES. *American Journal of Epidemiology*, 104(6):587– 592, 12 1976; and M. J. Knol and T. J. VanderWeele. Recommendations for presenting analyses of effect modification and interaction. *International Journal of Epidemiology*, 41(2):514–520, 2012

13 Carlo Berzuini and A. Philip Dawid. Stochastic mechanistic interaction. *Biometrika*, 103(1):89–102, 02 2016

14 https://www.R-project.org/

15 Giovanni Corrao, Federico Rea, Matteo Monzio Compagnoni, Luca Merlino, and Giuseppe Mancia. Protective effects of antihypertensive treatment in patients aged 85 years or older. *Journal of Hypertension*, 35(7):1432–1441, July 2017; and F Rea, G Occhino, A Cantarutti, et al. Is antihypertensive treatment protective in elderly frail patients? evidence from an italian real-world population. *Journal of Hypertension*, 37, 2019

16 Graziano Onder, Giovanni Rezza, and Silvio Brusaferro. Case-Fatality Rate and Characteristics of Patients Dying in Relation to COVID-19 in Italy. *Journal of the American Medical Association*, 323(18), May 2020; AB Docherty et al. Features of 20133 UK patients in hospital with Covid-19 using the ISARIC WHO clinical characterisation protocol: prospective observational cohort study. *BMJ*, 22, 2020; and S Richardson, JS Hirsch, M Narasimhan, et al. Presenting Characteristics, Comorbidities, and Outcomes Among 5700 Patients Hospitalized With COVID-19 in the New York City Area

17 Y Liu, Y Yang, C Zhang, et al. Clinical and biochemical indexes from 2019-nCoV infected patients linked to viral loads and lung injury. *Sci China Life Sci*, 2020

18 MR Deshotels, H Xia, S Sriramula, E Lazartigues, and CM Filipeanu. Angiotensin II mediates angiotensin converting enzyme type 2 internalization and degradation through an angiotensin II type I receptor-dependent mechanism. *Hypertension*, 64:1368–1375, 2014

19 CM Ferrario et al. Effect of angiotensin-converting enzyme inhibition and angiotensin II receptor blockers on cardiac angiotensin-converting enzyme 2. *Circulation*, 111:2605–2610, 2005

20 TE Walters, JM Kalman, SK Patel, M Mearns, E Velkoska, and LM Burrell. Angiotensin converting enzyme 2 activity and human atrial fibrillation: increased plasma angiotensin converting enzyme 2 activity is associated with atrial fibrillation and more advanced left atrial structural remodelling. *Europace*, 19:1280–1287, 2017; and

21 CA Dezsi and V Szentes. Effects of angiotensin-converting enzyme inhibitors and angiotensin receptor blockers on pro-thrombotic processes and myocardial infarction risk. *Am J Cardiovasc Drugs*, 16:399–406, 2016; BM Henry, J Vikse, S Benoit, EJ Favaloro, and G Lippi. Hyperinflammation and derangement of renin–angiotensin–aldosterone system in COVID-19: a novel hypothesis for clinically suspected hypercoagulopathy and microvascular immunothrombosis. *Clin Chim Acta*; and B Bikdeli, MV Madhavan, D Jimenez, et al. COVID-19 and thrombotic or thromboembolic disease: implications for prevention, antithrombotic therapy, and follow-up. *J Am Coll Cardiol*, 2020

22 R Fosboel et al. Association of angiotensin-converting enzyme inhibitor or angiotensin receptor blocker use with COVID-19 diagnosis and mortality. *JAMA*, 2020

23 G Mancia, F Rea, M Ludergnani, G Apolone, and G. Corrao. Renin-Angiotensin-Aldosterone System Blockers and the Risk of Covid-19. *N Engl J Med*., 2020

24 RD Lopes et al. Continuing versus suspending angiotensin-converting enzyme inhibitors and angiotensin receptor blockers: Impact on adverse outcomes in hospitalized patients with severe acute respiratory syndrome coronavirus 2 (SARS-CoV-2)– The BRACE CORONA Trial. *Am Heart J*., 226:49–59, 2020

